# The Healthcare Access and Delivery Survey: Turkiye Earthquake response

**DOI:** 10.1101/2023.04.22.23288485

**Authors:** Najeeb Rahman, İlker İnanç Balkan

## Abstract

**Background:** Conducting rapid health assessments during large-scale disasters can present significant challenges. While responding to key humanitarian needs is a priority, assessing and understanding gaps and areas for strengthening are vital in ensuring a comprehensive and equitable response.

**Aim:** To utilise the perspectives of healthcare workers responding to the Turkiye earthquake 2023, as a means of assessing humanitarian health response gaps.

**Design and setting:** Observational, descriptive, prospective study design in Turkiye.

**Method:** Online survey tool using a Likert-type scale, aligned with minimum standards of humanitarian health response themes, in both English and Turkish distributed by healthcare organisations and academic networks to a convenience sample of health workers responding to the earthquake.

**Results:** 45 respondents with perspectives demonstrating general limitations across all humanitarian health themes, with particular gaps in:

- Coordination and response timeliness
- Reporting and surveillance
- Mental health, palliative care and sexual and reproductive health

**Conclusion:** Using a survey tool to assess health worker perspective is feasible and provides rapid insights into humanitarian response efforts. Such data can be added to other assessment information to build a more comprehensive picture of needs while engaging with a key stakeholder group, and can support service providers to address gaps and strengthen response actions.

## Introduction

A 7.8 magnitude earthquake struck parts of central and southern of Turkiye as well as areas of north-western Syria ^1^. One of the challenges in any such large-scale disaster is in conducting rapid needs assessments to help guide response activities^2^. A further key consideration is the importance of prioritising, coordinating, and delivering relevant health services to meet the needs of communities and limit the disaster and displacement impacts on mortality and morbidity^3^. One approach to supporting assessments is to utilise health workers as stakeholders in generating information and insights on gaps and needs in response ^4^. The primary objective of this study was to support the health service development of Yeryuzu Doktorlari (YYD: Doctors Worldwide – Turkiye), a locally registered international medical charity, whose staff and volunteers were providing a medical response to earthquake victims. The secondary objective was to gain a wider view of health practitioners who were additionally involved in the response through other agencies.

## Method

A survey tool was developed based upon relevant sections of the Sphere Handbook: Humanitarian Charter and Minimum Standards in Humanitarian Response and can be found in Appendix A^5^. Statements were phrased in way which aligned to a) Core Humanitarian Principles, b) the Health System, and c) Health Services, as outlined in the handbook. The survey had previously been deployed in the UK amongst health workers in mid-2020 during the COVID-19 pandemic, and yielded important insights which although not formally published, were shared with local health leaders and managers to support plans.

Survey statements were initially drafted in English and then translated to Turkish with the assistance of one of the study authors, a Turkish first-language speaker and senior physician with experience of humanitarian contexts. A Likert-Scale approach was adopted where 1 represented Strongly Disagree, and 5 Strongly Agree, with 0 as a Don’t Know option.

A convenience sampling approach was used, with distribution of the online survey (using GoogleForms, Google, California, USA) via professional networks affiliated with Istanbul University Cerrahpaşa, as well as Yeryuzu Doktorlari. The survey was additionally shared with contacts at the Turkish Red Crescent Academy. Informed consent was obtained prior to collection of responses. Responses were collected for 10 days between the 24^th^ of February and 5^th^ of March 2023.

Basic descriptive analysis was performed to calculate mean scores for each question, which in turn was used to determine a domain average for each of the sections. Responses were further analysed by key demographics and cross-tabulations to observe for any trends, while acknowledging this was not a statistically powered survey given the nature of approach. In addition, free text comments were reviewed for relevance and qualitative narrative.

As this was considered a service development study, formal ethics review was deemed not required following the decision support tool by the NHS Health Research Authority^6^.

## Results

After screening for duplicates and inappropriate entries (eg: no relevant demographic details provided, or not related to this disaster), there were 45 unique respondents. The cumulative mean score for each domain is presented in Table 1 in relation to the total cohort as well as when adjusted by key demographic category. Tables 2-4 outline the average score of the total cohort against each of the domain statements to highlight priority areas. Only six comments of relevance were provided, which are presented in Table 5 along with the corresponding participant entry number.

**Table 1:**
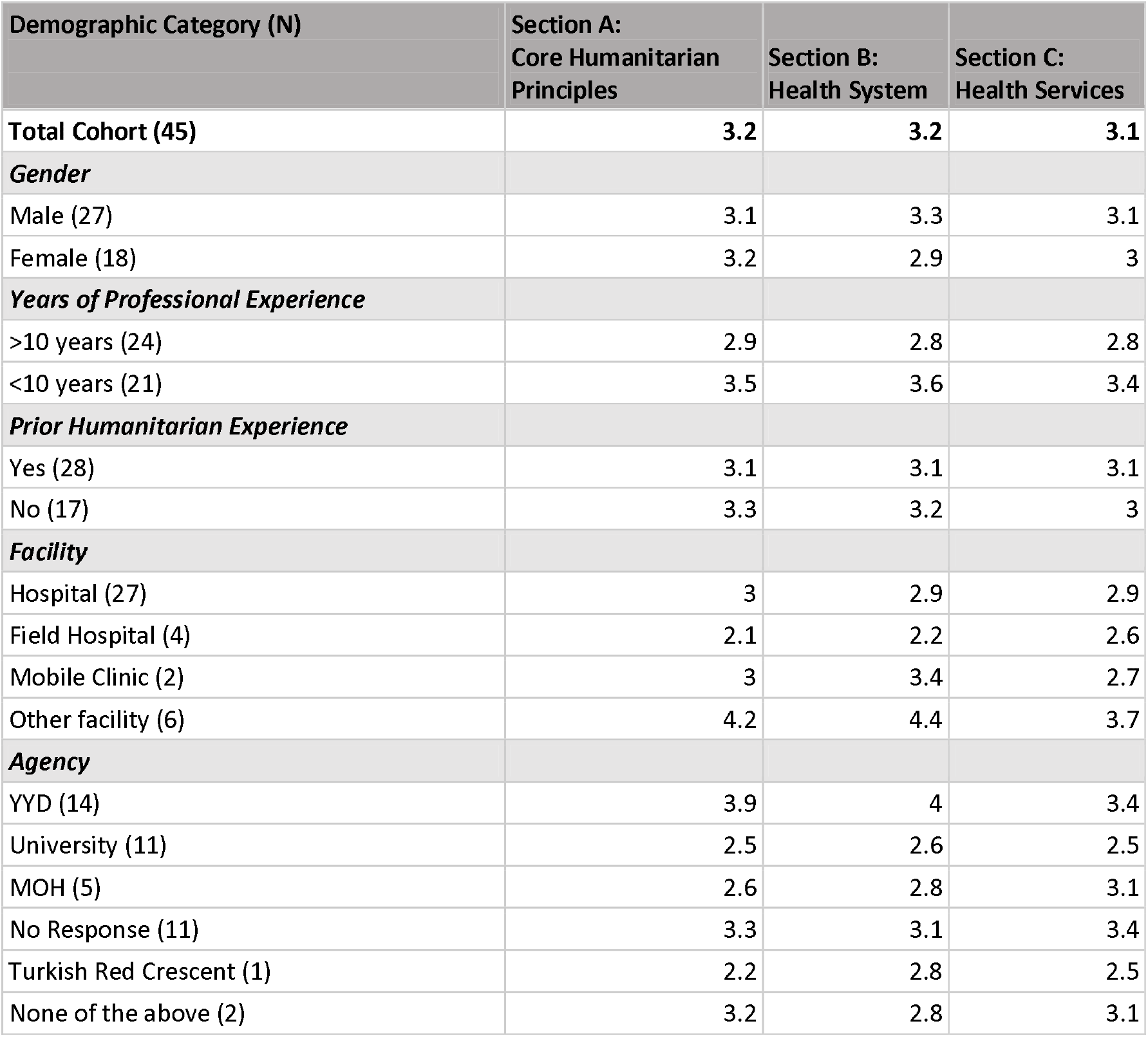
Summary of cohort demographic details as well as means scores in relation to the key humanitarian domains assessed.

| Demographic Category (N) | Section A:<br>Core Humanitarian<br>Principles | Section B:<br>Health System | Section C:<br>Health Services |
| --- | --- | --- | --- |
| **Total Cohort (45)** | **3.2** | **3.2** | **3.1** |
| **Gender** |  |  |  |
| Male (27) | 3.1 | 3.3 | 3.1 |
| Female (18) | 3.2 | 2.9 | 3 |
| **Years of Professional Experience** |  |  |  |
| >10 years (24) | 2.9 | 2.8 | 2.8 |
| <10 years (21) | 3.5 | 3.6 | 3.4 |
| **Prior Humanitarian Experience** |  |  |  |
| Yes (28) | 3.1 | 3.1 | 3.1 |
| No (17) | 3.3 | 3.2 | 3 |
| **Facility** |  |  |  |
| Hospital (27) | 3 | 2.9 | 2.9 |
| Field Hospital (4) | 2.1 | 2.2 | 2.6 |
| Mobile Clinic (2) | 3 | 3.4 | 2.7 |
| Other facility (6) | 4.2 | 4.4 | 3.7 |
| **Agency** |  |  |  |
| YYD (14) | 3.9 | 4 | 3.4 |
| University (11) | 2.5 | 2.6 | 2.5 |
| MOH (5) | 2.6 | 2.8 | 3.1 |
| No Response (11) | 3.3 | 3.1 | 3.4 |
| Turkish Red Crescent (1) | 2.2 | 2.8 | 2.5 |
| None of the above (2) | 3.2 | 2.8 | 3.1 |

**Table 2:**
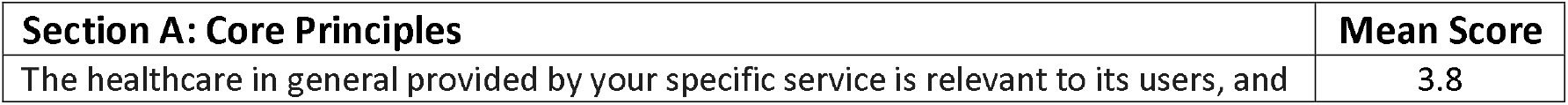
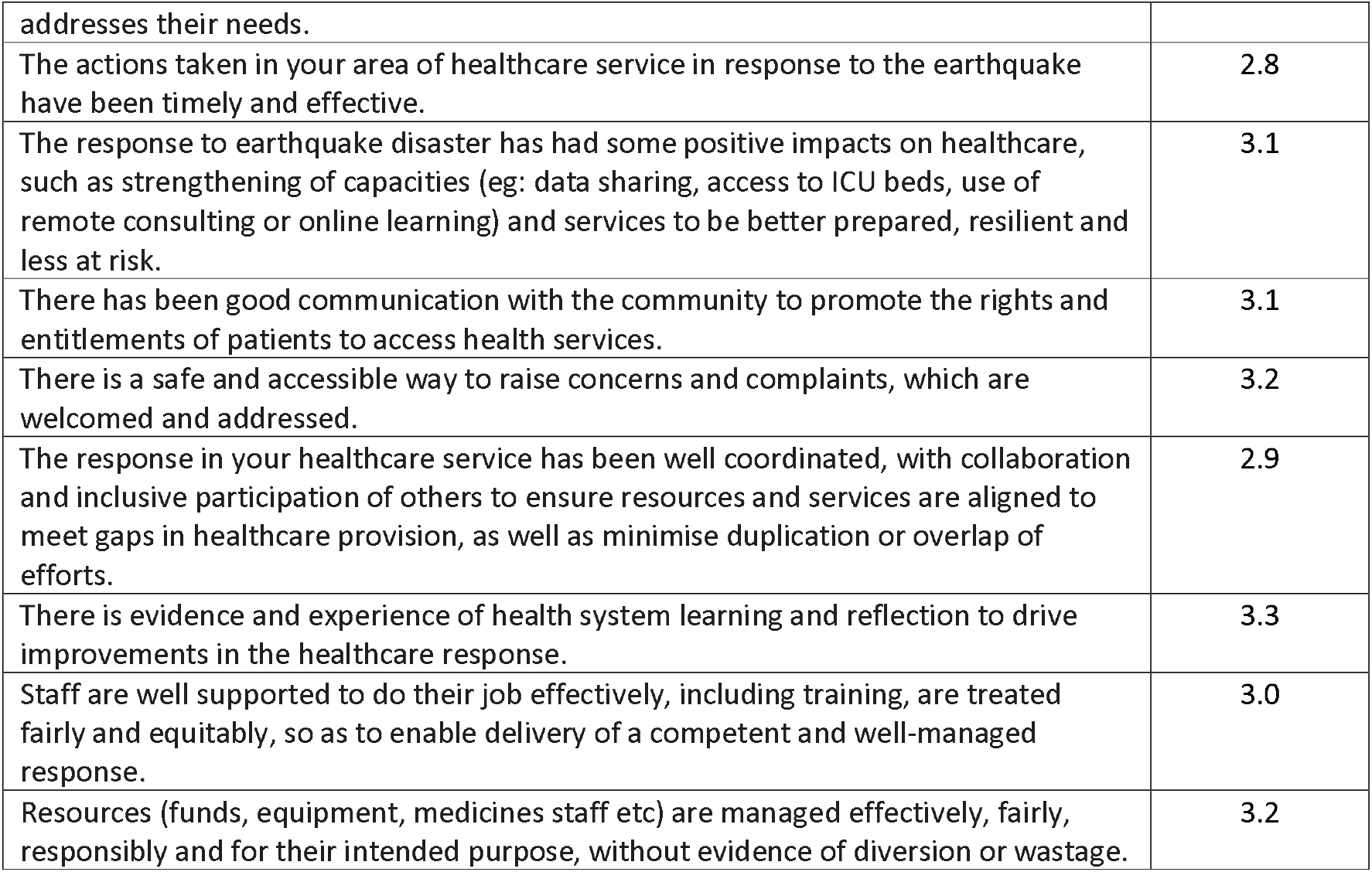
Summary of average scores of the total respondent cohort in relation to the section on core humanitarian principles aligned to health, covering themes of relevance, timeliness, capacity strengthening, promotion of rights and complaints management, coordination and collaboration, learning and improvement, workforce support and resource management.

**Table 3:**
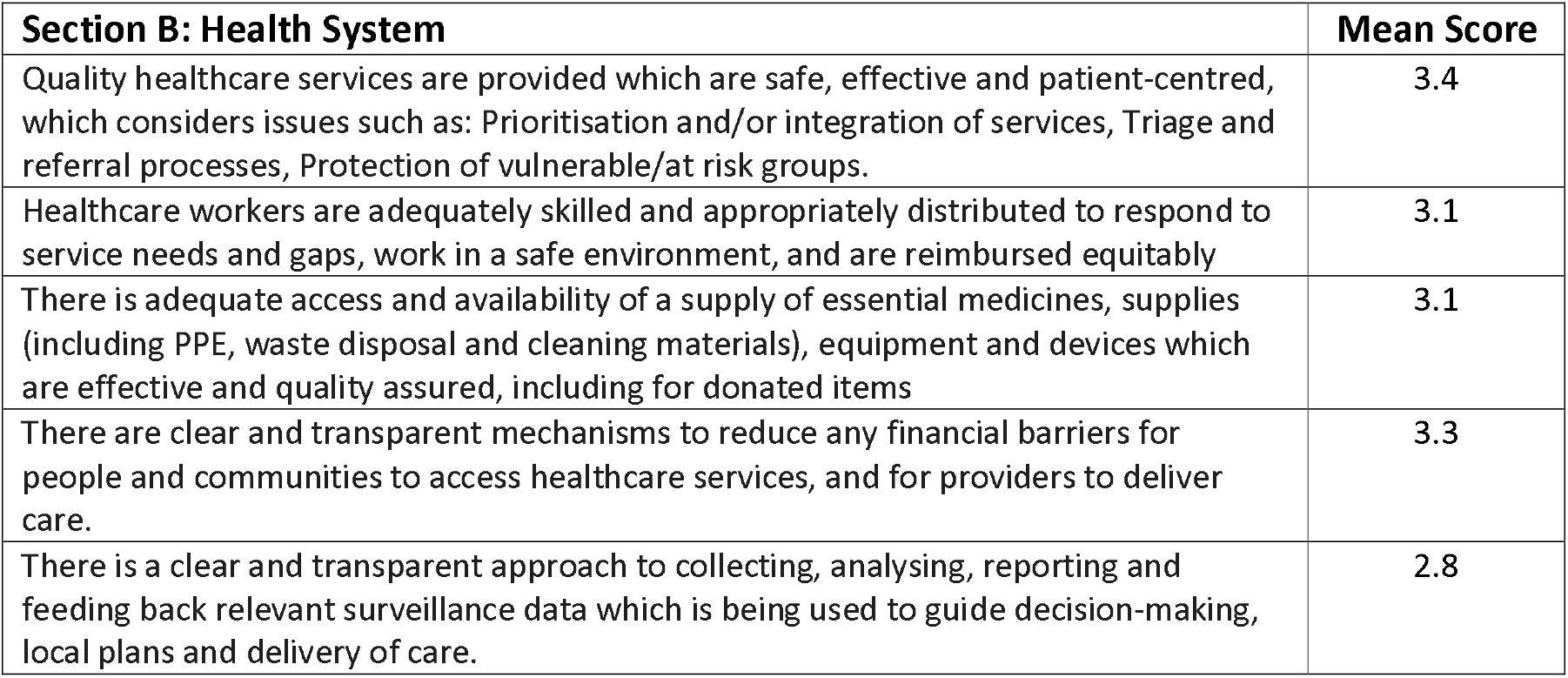
Summary of average scores of the total respondent cohort in relation to the section on health systems which includes themes of Quality, Human Resources, Supplies, Finance and Data.

**Table 4:**
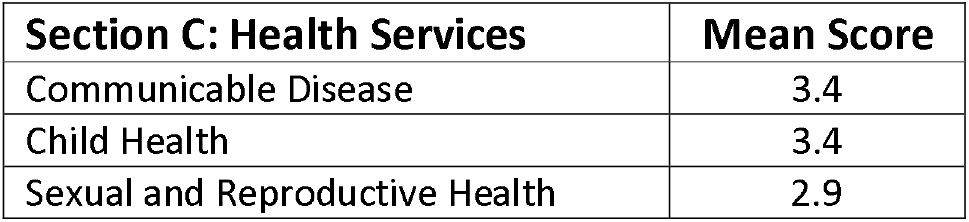
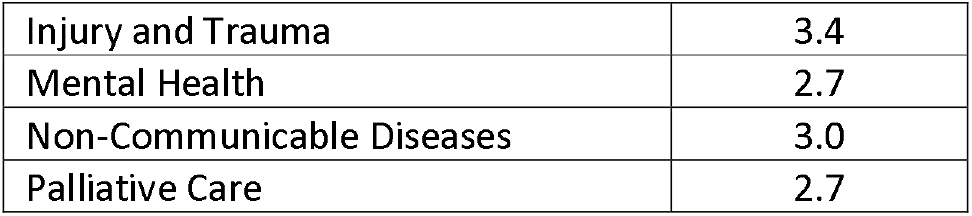
Summary of average scores of the total respondent cohort in relation to the section on essential health services.

| **Section C: Health Services** | **Mean Score** |
| --- | --- |
| Communicable Disease | 3.4 |
| Child Health | 3.4 |
| Sexual and Reproductive Health | 2.9 |
| Injury and Trauma | 3.4 |
| Mental Health | 2.7 |
| Non-Communicable Diseases | 3.0 |
| Palliative Care | 2.7 |

**Table 5:**
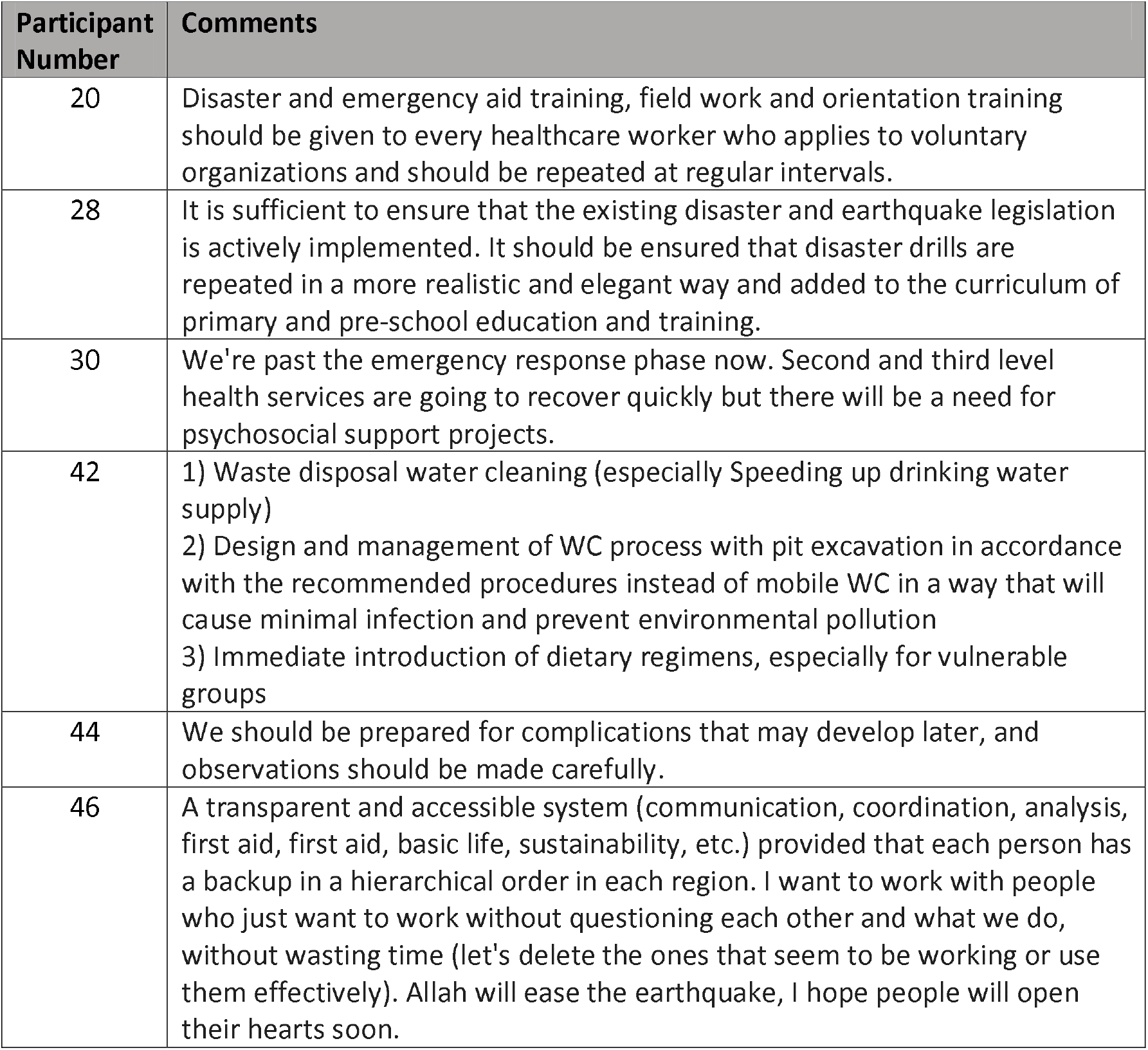
Summary the relevant narrative comments provided in relation to all 3 domains.

| Participant Number | Comments |
| --- | --- |
| 20 | Disaster and emergency aid training, field work and orientation training should be given to every healthcare worker who applies to voluntary organizations and should be repeated at regular intervals. |
| 28 | It is sufficient to ensure that the existing disaster and earthquake legislation is actively implemented. It should be ensured that disaster drills are repeated in a more realistic and elegant way and added to the curriculum of primary and pre-school education and training. |
| 30 | We're past the emergency response phase now. Second and third level health services are going to recover quickly but there will be a need for psychosocial support projects. |
| 42 | 1) Waste disposal water cleaning (especially Speeding up drinking water supply)<br>2) Design and management of WC process with pit excavation in accordance with the recommended procedures instead of mobile WC in a way that will cause minimal infection and prevent environmental pollution<br>3) Immediate introduction of dietary regimens, especially for vulnerable groups |
| 44 | We should be prepared for complications that may develop later, and observations should be made carefully. |
| 46 | A transparent and accessible system (communication, coordination, analysis, first aid, first aid, basic life, sustainability, etc.) provided that each person has a backup in a hierarchical order in each region. I want to work with people who just want to work without questioning each other and what we do, without wasting time (let's delete the ones that seem to be working or use them effectively). Allah will ease the earthquake, I hope people will open their hearts soon. |

## Discussion

There was good representation of participants across a range of experience, including those who volunteer with Yeryuzu Doktorlari, which represented the largest number of respondents (31%) who identified an affiliated agency. There was only one respondent affiliated with the Turkish Red Crescent. Similarly, most respondents were based at a hospital facility (60%) as opposed to mobile clinics (4%) or field hospitals (9%).

Given that the survey statements were mapped towards minimum standards of response, it can be suggested that in an ideal disaster response scenario, mean scores would be nearing 5, representing positive agreement with the principles and themes outlined. As such, where scores are less positively skewed, or indeed less than 3, it can be inferred that these are areas where gaps and needs are highlighted. Given that across the total 45 respondents the mean scores were only just above 3 for each of the domains suggests that the cohort viewed the response had areas where action could be improved and strengthened.

### Professional Experience

When exploring this in greater detail, and while acknowledging that this was not a study powered for statistical significance, it can be observed that respondents had a more negatively skewed perspective if they had more than 10 years of professional experience (scoring and average of 2.8 across all three domains) compared with those how had less than 10 years of experience (averaging 3.5).

### Agency affiliation

Similar trends were found if respondents were affiliated with a university teaching hospital, scoring an average of 2.5 across all three domains. This was not observed in respondents affiliated with YYD, where scores were skewed more positively towards an average of 3.8. Several participants (11) chose not to specify and agency and left this section empty. However, scores for this group in general trended positively.

### Facility

When cross tabulating facility of work with agency of affiliation, there was a distribution of representation, meaning it was not one group of agency respondents all working in the same facility type. Those working in hospitals had a near ambivalent perspective on the themes explored averaging 2.9. While the number of respondents working in field hospitals was limited (n=4), their views were negative with scores of 2.3, compared with any of the other facility categories.

### Prior humanitarian experience

Interestingly there was no large observed variation in perspective when comparing those with or without prior humanitarian experience, with scores of 3.1 and 3.2 respectively across all domains. When cross tabulating prior humanitarian experience with agency affiliation, it was observed that those based at university facilities with previous experience in general had a more negative view of the response, but a confounding factor was that all those participants also had greater than 10 years of professional experience. This grouping was not observed in other cross tabulation, and the averages scores were additionally not as negatively skewed.

### Domain review

While cumulative means across the total cohort for each domain has been presented in Table 1. Tables 2-4 provide a breakdown of scores against each statement of the themed domain. Reviewing these scores provides further insights on strengths and weaknesses of the response. The cohort was positive that the general healthcare provided was relevant and specific to needs, with a score of 3.8, which considering core humanitarian principles.

However, when reviewing key aspects of essential health service delivery (Table 3), mental health and palliative care were areas of deficiency, with scores of 2.7 each. While not as negatively skewed, sexual and reproductive health as well as non-communicable disease services were also felt not to be as comprehensively delivered (scoring 2.9 and 3.0 respectively) as other health areas. Further key areas representing an opportunity to strengthen activities when considering both principles of humanitarian response, as well as the health system related to timeliness, coordination and data management. The statements provided highlight similar areas of concern.

## Limitations

While this study sought to be easily accessible through wide distribution networks, response rates were limited. This may have been due to how occupied people were in relation to the response, or due to the nature of themes explored or who was conducting the study, and whether potential participants felt willing and able to respond and trust the nature of the study given its opportunistic approach. The survey was kept open for a short period of 10 days, and so it is unclear how effective promotion and distribution would have been during this time and at the early stage of the response. In addition, although a senior Turkish academic was involved in this project, it was initially designed in English and on humanitarian themes which have originated initially in Western discourse. As such, there may have been some contextual and cultural issues which may have additional contributed to differential expectations on survey participation and completion. Further consideration needs to be given to whether face-to-face collection would improve response rates, but this would require additional resource and planning. Inferences on specific areas highlighted should be interpreted with an understanding of the smaller cohort. In addition, despite a range of perspectives, comments were only provided by a minority of respondents to help explain some of their views. This may again reflect on the issues highlighted regarding culture and expectations on what the study was about.

## Conclusions

From a service development perspective of those managing a response to a large-scale crisis or disaster, the approach presented provides rapid insights into key standardised humanitarian and health themes. The tool is easy to administer and provides views from an important stakeholder group (health workers) involved in delivering services, and so could serve as a further adjunct to community needs assessment process, as well as highlight priority areas. The study provided insights that the overall response could be strengthened, but with focus on coordination, data management and timeliness. Mental health and palliative care services were specifically highlighted as areas to improve, and where further in-depth assessments are indicated to support programming to meet community and stakeholder needs. Additional exploration is warranted (eg: through focus group discussions or participant interviews) given the wide variation of perspectives between those with greater than 10 years of professional experience compared with more junior staff and volunteers. Further work is needed to better understand and advance the utility and validity of this survey as a standardised approach to rapid, pragmatic insight development which could be embedded as part of a Monitoring, Evaluation, Accountability and Learning (MEAL) approach in humanitarian response contexts. This study also demonstrates the need for agile research and service evaluation processes which require collaboration, investment and innovation to accompany response delivery.

## Supporting information

Appendix A: Outline of information for participants as well as Survey Tool

## Data Availability

Data produced in this study are available upon reasonable request to the authors

## Acknowledgements

The authors would like to extend our thanks and appreciation to the staff at Yeryuzu Doktorlari as well as Turkish Red Crescent for supporting the distribution of this survey at a time of intense activity.

We would also like to thank Dr Mohammed Owais Rahman, who supported initial development of the online GoogleForm, Dr Ali Ihsan Nergiz who supported translation of the narrative comments, as well as Dr Saraah Khalid who assisted in converting the agreement responses to numerical values.

