## Appendix A: Outline of information for participants as well as Survey Tool for "The Healthcare Access and Delivery Survey: Turkiye Earthquake response"

*NB: Information was translated and provided to participants in Turkish language as standard. These materials can be provided by corresponding with the author*. *Statements in sections 2-4 were appraised against a 5-point Likert scale from Strongly disagree to Strongly agree, with a separate ‘Don’t know’ option*

**Section 1: Information for Participants: What is this assessment about?**

It is recognised that there have been significant challenges in the delivery of healthcare services in response to earthquake disasters. The humanitarian sector has learned approaches to support the strengthening of health care provision by focusing on key principles, systems and services. By utilising a framework derived from the humanitarian health sector, this assessment seeks to rapidly evaluate key themes and areas that would benefit from further focus and improvement. If you are a front line health worker we are keen to hear your views.

**Who is carrying out the survey?**

The primary investigator is Dr Najeeb Rahman, who is a Trustee of Doctors Worldwide UK, a humanitarian practitioner, and an Emergency Medicine consultant based at Leeds Teaching Hospitals NHS Trust, UK. This survey is being conducted on behalf of Doctors Worldwide, and where findings may be shared in the public domain through reports and publications.

**What is involved?**

We ask that you complete this short survey with regard to your opinions and experiences on the current health service response to the earthquake in Turkiye We ask that you complete it honestly. The survey will take around 5-10 minutes to complete.

Thank you for taking part in this assessment, and helping to improve our understanding and response to the challenge of the earthquake in Turkiye. This would not be possible without your support and involvement.

**Section 2: Core response principles**

Please select your agreement or disagreement which most accurately reflects your opinion on the statement below.

1. The healthcare in general provided by your specific service is relevant to its users, and addresses their needs.
2. The actions taken in your area of healthcare service in response to the earthquake have been timely and effective.
3. The response to earthquake disaster has had some positive impacts on healthcare, such as strengthening of capacities (eg: data sharing, access to ICU beds, use of remote consulting or online learning) and services to be better prepared, resilient and less at risk.
4. There has been good communication with the community to promote the rights and entitlements of patients to access health services
5. There is a safe and accessible way to raise concerns and complaints, which are welcomed and addressed.
6. The response in your healthcare service has been well coordinated, with collaboration and inclusive participation of others to ensure resources and services are aligned to meet gaps in healthcare provision, as well as minimise duplication or overlap of efforts.
7. There is evidence and experience of health system learning and reflection to drive improvements in the healthcare response.
8. Staff are well supported to do their job effectively, including training, are treated fairly and equitably, so as to enable delivery of a competent and well-managed response.
9. Resources (funds, equipment, medicines staff etc) are managed effectively, fairly, responsibly and for their intended purpose, without evidence of diversion or wastage.
10. Do you have any further comments or feedback on the questions in this section which relate to the humanitarian response in general?

**Section 3: The health system**

11. Quality healthcare services are provided which are safe, effective and patient-centred, which considers issues such as: Prioritisation and/or integration of servicesTriage and referral processes Protection of vulnerable/at risk groups.
12. Healthcare workers are adequately skilled and appropriately distributed to respond to service needs and gaps, work in a safe environment, and are reimbursed equitably.
13. There is adequate access and availability of a supply of essential medicines, supplies (including PPE, waste disposal and cleaning materials), equipment and devices which are effective and quality assured, including for donated items.
14. There are clear and transparent mechanisms to reduce any financial barriers for people and communities to access healthcare services, and for providers to deliver care.
15. There is a clear and transparent approach to collecting, analysing, reporting and feeding back relevant surveillance data which is being used to guide decision-making, local plans and delivery of care.

**Section 4: Essential healthcare**

16. Communicable Diseases: Information on the prevention, surveillance, case management, and referral of communicable diseases is available and accessible to staff. (Consider conditions such as measles, respiratory illnesses, diarrhoeal illnesses, meningitis, chest infections and sexual transmitted illnesses, vaccine-preventable illnesses, urine infections)
17. Communicable Diseases: Care for acute infections such as soft tissue infections/cellulitis urinary tract infections, chest infections, as well as complications such as suspected sepsis is timely, effective, culturally competent and available to all groups of patients.
18. Child Health: Essential services such as vaccinations, safeguarding surveillance newborn and child health checks are being accessed adequately, and delivered safely and effectively.
19. Child Health: Care for acute illness and injury, including febrile illnesses is timely, effective, culturally competent and available to all groups of patients.
20. Sexual and Reproductive Health: Essential services such as family planning and antenatal care, including termination of pregnancy, emergency contraception, HIV post exposure prophylaxis, and management of STIs are being accessed adequately, and delivered safely and effectively.
21. Sexual and Reproductive Health: Care for complications of pregnancy such as miscarriages, complicated deliveries, hyperemesis, gestational diabetes, hypertension is timely, effective, culturally competent and available to all groups of patients.
22. Injury and Trauma: Essential services such as access to X-Ray, assessment, management and follow-up of fractures and soft tissue injuries are being accessed adequately, and delivered safely and effectively.
23. Injury and Trauma: Care of injuries related to crush and other accidents is timely, effective, and available to all groups of patients (including prehospital, surgical, critical care, radiology and rehab).
24. Mental Health: Essential services such as community-based and specialised mental health assessments, management of acute stress or depression, post-traumatic stress, medicines prescriptions are being accessed adequately, and delivered safely and effectively.
25. Mental Health: Care of serious mental health illness, self-harm, and harm related to alcohol and drugs is timely, effective, culturally competent and available to all groups of patients.
26. Non-Communicable Diseases: Essential services to support screening, prevention and management of longer-term care of diseases such as cardiovascular disease, diabetes, asthma, COPD, and cancer care are being accessed adequately, and delivered safely and effectively.
27. Non-Communicable Diseases: Care of acute complications of diseases such as Myocardial Infarction, Stroke, exacerbations of chronic lung disease, or complications of cancer is timely, effective, culturally competent and available to all groups of patients.
28. Palliative Care: Essential services to maximise symptom control, comfort, dignity and quality of life, and end of life care for patients, in addition to support for family members, are being accessed adequately, and delivered safely and effectively.
29. Palliative Care: Care planning and provision or referrals of acute support for patients who are dying from earthquake trauma and non-trauma related causes is timely, effective, culturally competent and available to all groups of patients.

**Section 5: About You**

30. What is your age?
31. What is your gender?
32. What is your profession?
33. How many years of professional practice do you have?
34. Do you have any prior experience in humanitarian response?
35. Where are you currently working (location) City, town, district ?
36. What kind of facility do you currently working in?

- Hospital
- Field Hospital
- Mobile clinic
- Primary care clinic

37. Which organisation do you work for?

- Ministry of Health (including university hospitals)
- AFAD
- Red Crescent
- DWWT
- Other

38. Do you have any further comments, feedback, suggestions, or anything else to add which may assist with the direction of the emergency medical response for the Earthquake survivors?
